# Sodium glucose co-transport inhibitors to treat heart failure in patients with complex adult congenital heart disease - a systematic review and meta-analysis

**DOI:** 10.64898/2026.01.14.26344158

**Authors:** Rachel M. Wald, Nili Schamroth Pravda, Jasmine Grewal, S. Lucy Roche, Rafael Alonso-Gonzalez, Jacob A. Udell, Candice K. Silversides, Hwee Teoh, Adrian Quan, C. David Mazer, Subodh Verma, George Tomlinson, Anoop S V Shah

**Affiliations:** Peter Munk Cardiac Centre, University Health Network, University of Toronto, Toronto, ON, Canada; Hadassah Medical Center, Jerusalem, Israel; Toronto General Hospital Research Institute, Toronto, ON, Canada; Tel Aviv Medical Center Ichilov, Gray Faculty of Medical and Health Sciences, Tel Aviv University, Tel Aviv, Israel; Providence Health Centre, University of British Columbia, Vancouver, BC, Canada; Women’s College Hospital, University of Toronto, Toronto, ON, Canada; St. Michael’s Hospital, University of Toronto, Toronto, ON, Canada; London School of Hygiene and Tropical Medicine, London, England; Imperial College London National Health Service Trust

## Abstract

**Background:** Sodium glucose co-transport inhibitors (SGLT2i), although established heart failure (HF) therapy in acquired heart disease, are not well-studied in adult congenital heart disease (ACHD). We aimed to conduct a systematic review and meta-analysis of SGLT2i therapy in moderate or severe complexity ACHD.

**Methods:** Five databases (Pubmed, Medline, Embase, SCOPUS, and Cochrane) were searched for peer-reviewed journal articles describing SGLT2i HF therapy in moderate or severe complexity ACHD. Outcomes included adverse clinical events, biochemical markers of HF (N-terminal pro-brain natriuretic peptide [NT-proBNP] or BNP), and imaging markers of cardiac function (global longitudinal strain [GLS] and fractional area change [FAC]). Forest plots demonstrated mean study effects as individual and pooled estimates. The impact of heterogeneity on the overall variance was evaluated.

**Results:** The systematic review included 10 studies (n=174 patients, 60% male). SGLT2i therapy was associated with a statistically significant improvement in GLS (mean difference -1.6 [-2.4,-0.9]) but not FAC (mean difference +1.86 [-6.2,+9.9]); there was no significant post therapy change in NT-proBNP or BNP (mean difference -240 pg/mL [-516,45] and -52 pg/mL [-129,26], respectively). Heterogeneity for the pooled effects for GLS and FAC was low (I^2^=0%), although moderate to high for NT-proBNP and BNP (I^2^=47% and I^2^=90%, respectively). Data were insufficient for evaluation of SGLT2i impact on clinical outcomes.

**Conclusions:** Pooled results across studies suggest that SGLT2i therapy can improve GLS among people with ACHD-HF, however the clinical implications of this observation warrant further study. Randomized controlled trials are now needed to evaluate the impact of SGLT2i therapy in ACHD.

## INTRODUCTION

There is an expanding evidence base which unequivocably supports the pharmacologic treatment of HF in adults with acquired forms of heart disease.^1,2^ In stark contrast, supportive evidence to guide medical therapy to address heart failure (HF) in adult congenital heart disease (ACHD) is virtually absent. Sodium glucose co-transport inhibitors (SGLT2i) are among the newest and most promising medications for treatment of HF in acquired heart disease. This class of medications has had a profound impact on patients with acquired HF and is now considered a cornerstone in contemporary guideline directed medical therapy (GDMT). However, it remains unclear whether these drugs are also effective in ACHD.

The risk of HF in the ACHD population increases with age, with an inflection point noted in the fifth decade of life.^3^ In patients with complex ACHD, HF prevalence has been observed to be 14.8% at 42 years as compared with 1.5% in the general population.^4^ The risk is higher in patients with a systemic right ventricle (sRV) where HF prevalence reaches 25%, and is higher still in those with single ventricle (SV) physiology where prevalence rises to 30%. Despite the urgent need for effective medical therapies to address ACHD-HF, there is a glaring lack of evidence to guide contemporary management. While little is known about SGLT2i in the treatment of ACHD-HF in general as these patients are not featured in any of the large outcome trials, data are particularly lacking in patients with sRV or SV anatomies, the subset of patients at greatest risk of HF, with no robust randomized controlled trials published to date.

These knowledge gaps highlight the need for a systematic review and meta-analysis of the existing literature on the use of SGLT2i therapy to provide evidence-based HF management in this high-risk subgroup of ACHD patients. We hypothesized that there would be a beneficial effect of SGLT2i therapy on clinical outcomes as well as surrogate outcome measures of HF, defined as serum biomarkers, imaging measures of ventricular systolic function and exercise test parameters of functional capacity.

## METHODS

We aimed to synthesise available evidence evaluating the potential therapeutic role of SGLT2i in ACHD patients with HF and moderate or severe complexity disease. The main study objective was to evaluate the cardiovascular effects of SGLT2i in this population by conducting a systematic review of the available literature and performing a meta-analysis on suitable studies. Exploration of the safety profile of SGLT2i was a defined as a secondary objective.

### Inclusion and exclusion criteria

Articles were included if SGLT2i therapy was initiated to treat ACHD-HF in patients with complex lesions, specifically sRV anatomy or SV physiology, a quantifiable treatment effect (cardiovascular outcome) was provided (i.e. mean with standard deviation or median with range) and ≥5 patients were described. Only published, peer-reviewed journal articles in medical databases in the English language were included. Articles were excluded if a treatment effect could not be determined according to anatomic subtype (specifically sRV or SV) or patients were <18 years of age. The sRV classification included patients with complete transposition of the great arteries (D-TGA) following atrial redirection surgery (Mustard or Senning palliations) and the SV category included patients with univentricular physiology post Fontan palliation; patients with Eisenmenger physiology or unpalliated cyanotic forms of congenital heart disease (CHD) were excluded.

The PICOTS approach (Population, Intervention, Comparison, Outcome, Timing and Setting) was used. Specifically, the population was defined as patients with HF and complex ACHD consisting of sRV or SV and age ≥18 years. The intervention was restricted to treatment with SGLT2i therapy (empagliflozin, dapagliflozin and/or canagliflozin). The comparison to SGLT2i use was either placebo versus routine care or pre/post-intervention with SGLT2i (delta change). Outcomes of interest included clinical endpoints (death or hospital admission for an adverse cardiovascular event) or change in surrogate outcomes defined as serum biomarkers, imaging measures of systolic function, exercise testing parameters and/or patient reported outcomes (questionnaires). Timing was defined as ≥ 12 weeks of therapy with adequate follow-up. The setting of all studies was ambulatory (outpatient) care.

### Search strategy

Five databases (Pubmed, Medline, Embase, Scopus and Cochrane) were systemically searched using the search terms sodium glucose cotransporter 2 inhibitor, empagliflozin, dapagliflozin, canagliflozin, congenital heart disease, single ventricle, Fontan procedure, systemic right ventricle, Mustard surgery, Senning surgery, B-type or brain natriuretic peptide, exercise test, 6 minute walk distance or outcomes. Details of the full search strategy, which extended until September 2025, are shown (Supplementary Appendix).

### Data extraction

Data recorded included, but were not limited to: study characteristics (author, location of study, year of publication, setting and study design); population (sample size, participant characteristics [age, sex, anatomic lesion]); medications (including SGTL2i type and dose; other HF medications if listed); outcomes; duration of follow-up and loss to follow-up; and number of patients discontinuing therapy along with reasons why. Additionally, all reported drug-related side effects were categorized.

In studies describing a range of ACHD anatomies, data pertaining to the subsets of patients with sRV or SV were preferentially extracted for focused analysis. Similarly, for studies with a range of ages including both pediatric and adult patients, only data pertaining to adults ≥ 18 years of age were entered into the analysis. In select circumstances, patient-level outcome data were directly analyzed, if available and as necessary, in order to satisfy the inclusion criteria of the present study.

Study methodology, results and presentation are according to the Preferred Reporting Items for Systemic Reviews and Meta-Analysis (PRISMA) guidelines with a checklist provided (Supplementary Table 1). Risk of bias assessment was primarily evaluated using the Cochrane Collaboration Risk of Bias Tools for Non-Randomised Studies of Interventions or Randomized Controlled Trials (ROBINS-I or ROB-2 tools, respectively). As there is no specific risk of bias tool for pre/post-intervention studies, an additional tool was added for robustness of bias assessment, the Newcastle-Ottawa Scale (Supplementary Figures 2-4).^5,6^

Note was made of the use of concomitant medications which could enhance the effect of SGLT2i (i.e. other forms of GDMT) or which could conversely limit tolerability (ie diuretic use). Additionally, severity of HF at study outset was noted (New York Heart Association functional class) when available, as disease severity may limit response to SGLT2i therapy (patients who are relatively well at study entry may not have an observed response to treatment). Differential responses to SGLT2i therapy may exist according to patient subtype (the impact of SGLT2i may differ in patients with a biventricular circulation such as sRV as compared with patients with SV physiology), and this was recorded whenever possible.

The screening process that was employed is depicted in Figure 1. In total 414 studies were identified across 5 databases. Duplicate studies were then excluded. Two experienced ACHD physicians (RMW and NSP) independently screened all abstracts obtained by the systematic search and study eligibility was determined by consensus. A total of 17 abstracts which met the PICOTS criteria were selected for further full-text review. Ultimately, 10 studies were selected for inclusion in the systematic review having met PICOTS and inclusion critera.^7–16^

**Figure 1.**
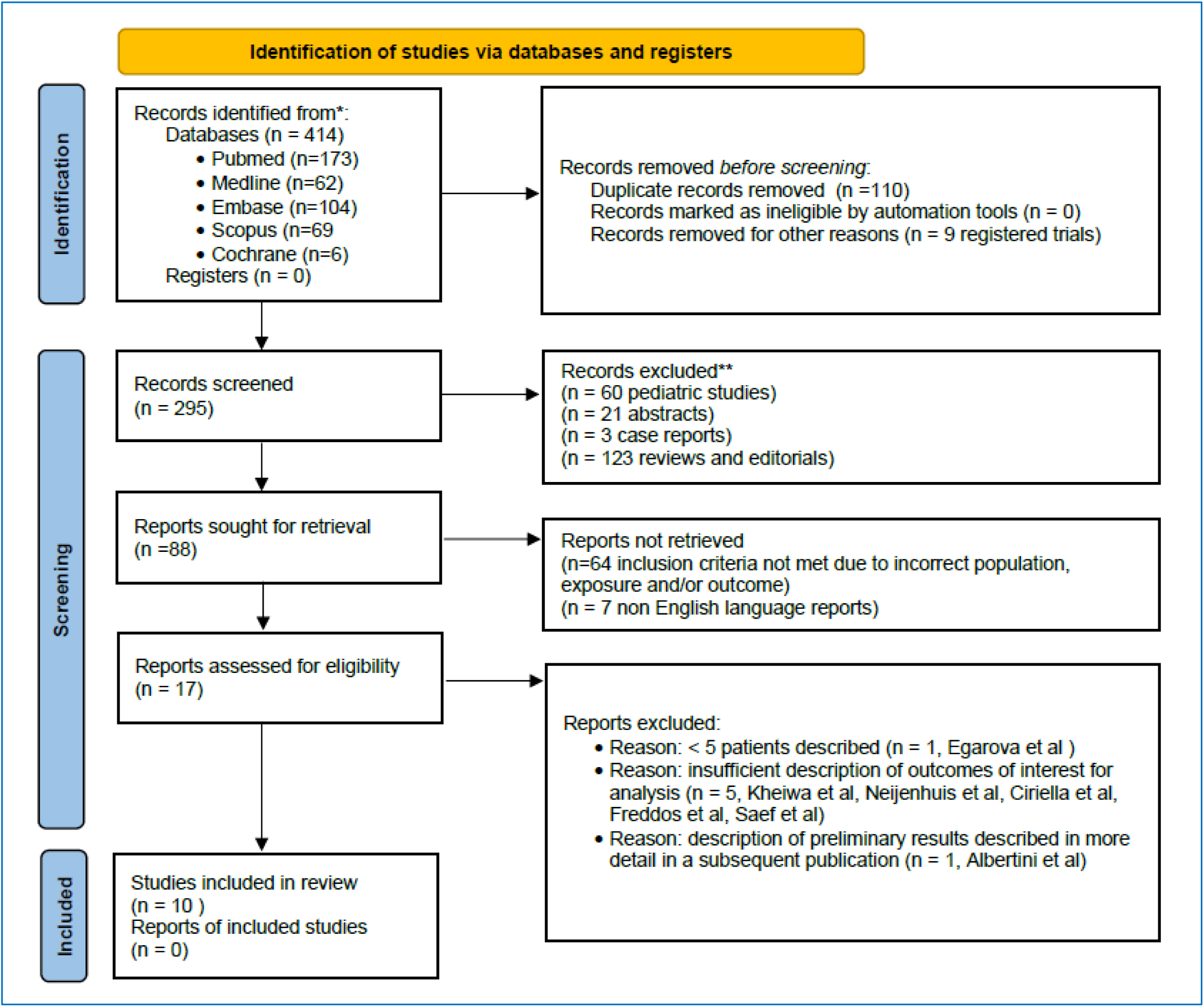
PRISMA Diagram Demonstrating Selection of Studies for Inclusion. Of the 17 studies assessed for eligibility, 7 were excluded^13,35–40^ (for reasons shown above) and 10 studies were ultimately included for systematic review^7–16^. *Consider, if feasible to do so, reporting the number of records identified from each database or register searched (rather than the total number across all databases/registers). **If automation tools were used, indicate how many records were excluded by a human and how many were excluded by automation tools.

### Outcomes

Measures of HF outcomes in this study encompassed multiple domains. These included clinical outcomes (major adverse cardiovascular events such as death, transplant, hospitalization for HF) and patient reported outcomes (using validated survey instruments). Because major adverse cardiovascular events may not be apparent in short or mid-term follow-up, surrogate outcomes were also identified for inclusion, specifically: (1) biochemical measures of HF (change in NTproBNP or BNP); (2) cardiovascular imaging measures of systolic function in the systemic ventricle on cardiovascular magnetic resonance imaging (CMR) or echocardiography including % ejection fraction (EF), % fractional area change (FAC) and/or % global longitudinal strain (GLS); and (3) exercise testing parameters (metres walked on a 6-minute walk test or peak oxygen consumption [VO_2_] as measured by mL/kg/min or % predicted on cardiopulmonary exercise testing). For assessment of ventricular function, CMR studies were preferentially selected ahead of echocardiography if available; only 3D-derived measures for EF on echocardiography were considered for analysis (although 3D EF on echocardiography is not validated in these patient subsets, 2D estimation of EF on echocardiography is considered even less reliable in the context of complex ventricular geometries such as sRV and SV).

### Statistical analysis

Data are presented as descriptive summaries in tabular or barplot format. Study data reported as median (range or IQR) were converted to mean (standard deviation), as previously reported.^17–20^ Both fixed and random effect estimates of treatments were calculated. Forest plots were used to graphically display mean study effects (individual and pooled estimates) and to visually examine heterogeneity between studies. Between study heterogeneity was presented as the I^2^ (the percentage of total variability between estimates attributable to intrinsic heterogeneity rather than sampling error) and assessed using the Cochran Q test. Risk of publication bias was explored with funnel plots and associated Egger tests. Statistical analysis was completed using R Core Team (2025 R Foundation for Statistical Computing, Vienna, Austria).

## RESULTS

### *S*tudy characteristics and patient population

Ten studies met inclusion criteria (Figure 1) and contributed a total of 174 adults with either sRV or SV anatomies (60% male, median reported age 37.5 years [range 21 to 54 years) (Table 1).^7–16^ The majority of the studies were cohort studies (80%, n=8); there was 1 open label randomized trial (with 25 cases and 25 controls) and 1 case series (detailing drug safety in 5 adults). In all studies, the effect was expressed as the mean or median difference in the outcome measure following treatment with SGLT2i within a patient (pre/post-treatment) or between patients (case versus control). All studies took place in tertiary care centres. Studies originated in Europe (50%, n=5), the United States (40%, n=4) and Japan (10%, n=1) (Supplementary Figure 1).

**Table 1:**
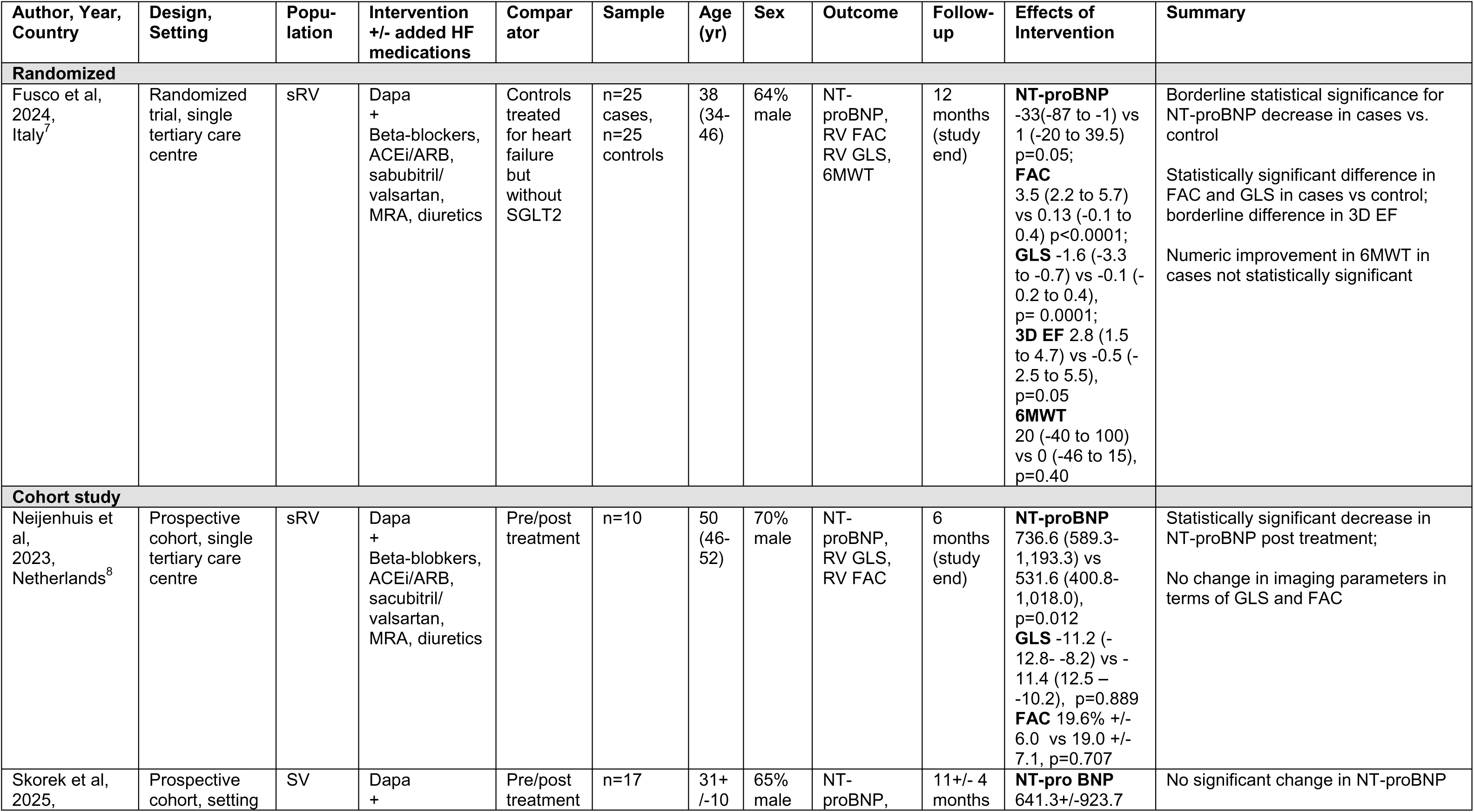
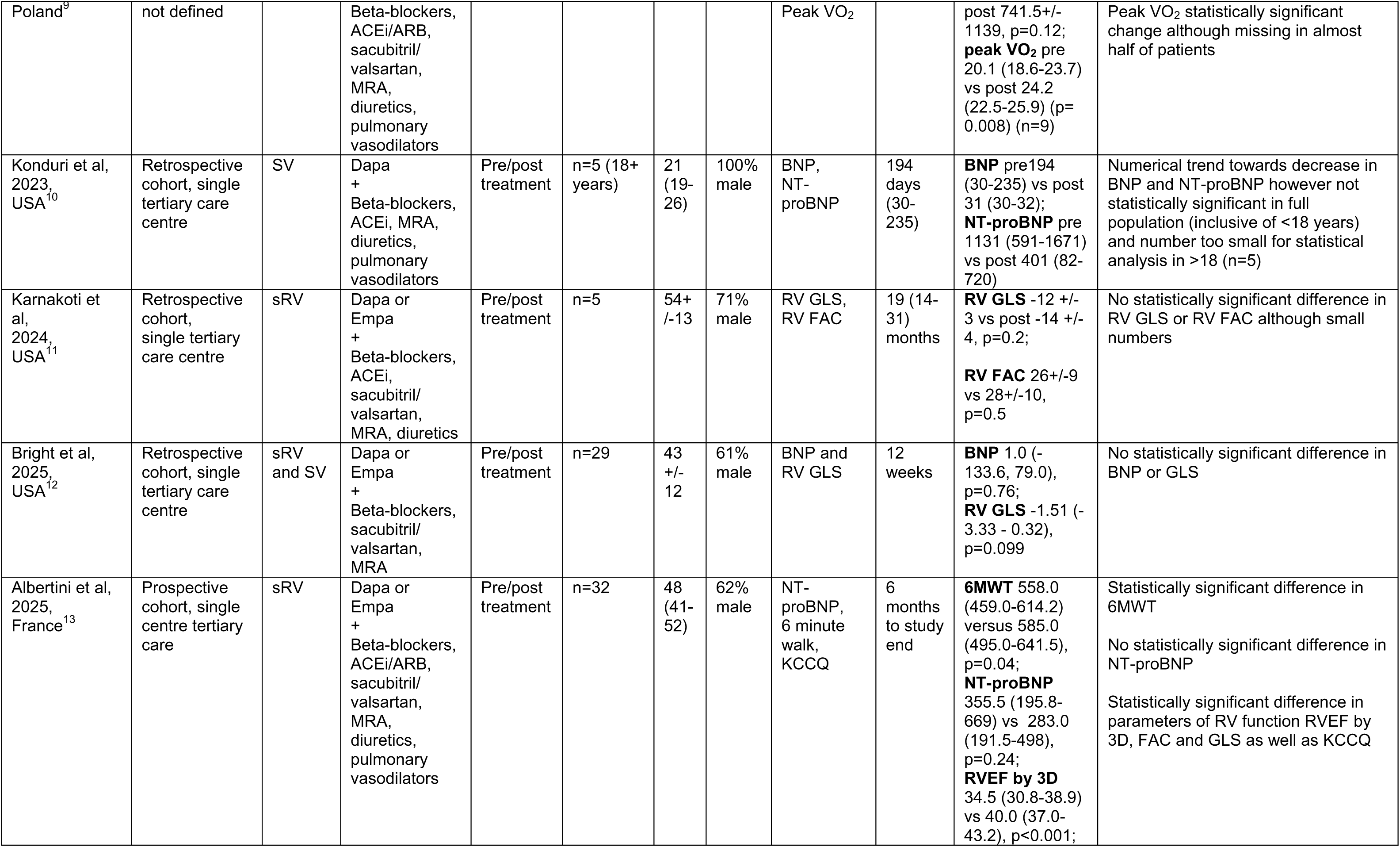
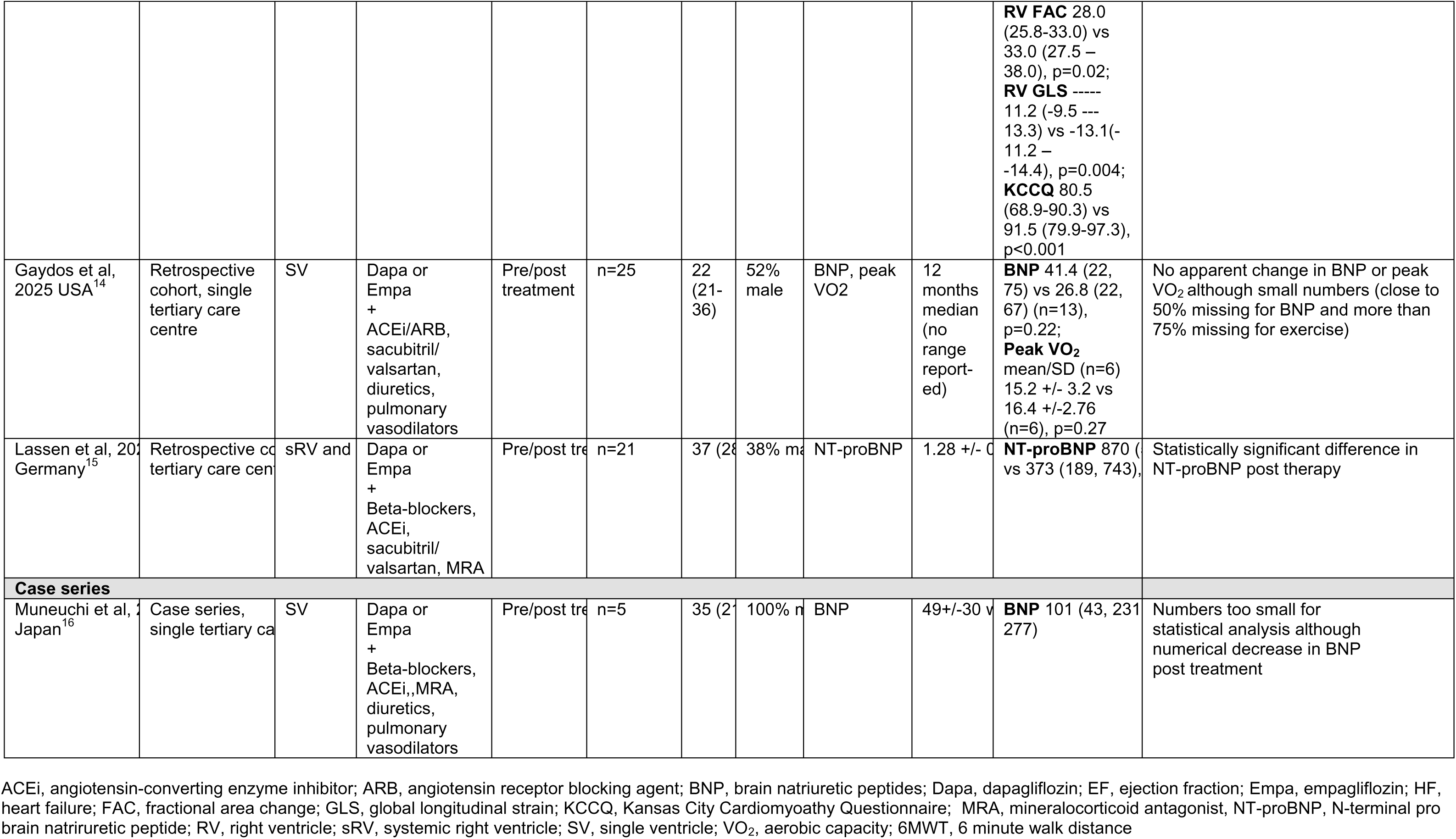
Studies Meeting Inclusion Criteria (n=10)

With respect to CHD anatomies, 4 studies reported exclusively on patients with underlying sRV and 4 studies reported only on patients with underlying SV. There were 2 studies which reported on both sRV and SV lesions, among other types of ACHD anatomies (Table 1). The largest study reported results from a prospective observational cohort of 32 patients^13^ and the smallest had 5 patients;^10,11,16^ 4 studies had 10 or fewer patients.^8,10,11,16^

### Intervention

Four studies (40%) described dapagliflozin use exclusively and 6 studies (60%) described the use of dapagliflozin and/or empagliflozin (Table 1). The dose of the SGT2i used was reported in 8 studies and not explicitly stated in 2 studies.^12^ ^9^ Patient adherence to medications as prescribed was only ascertained in one study through telephone contact with patients.^12^ The concomitant use of additional HF medications were noted in all studies, however only one study reported on whether patients were optimized on their HF therapies^12^ and no studies controlled for SGLT2i response with adjustment for receipt of other medications.

### Outcomes

Outcome measures are listed in Table 1 along with the corresponding effect of the related intervention. Data on adverse clinical outcomes were insufficient for analysis (one case series reported on the absence of unexpected hospitalizations following SGLT2i therapy in the 5 patients described^16^ and in one cohort study the frequency of HF related hospitalization was decreased from 4 admissions in the 6 months prior to therapy to 1 admission in the 6 months following therapy^8^). The main measures selected for this systematic review were serum biomarkers (NT-proBNP and BNP), imaging measures of ventricular function (FAC, GLS and 3D EF) and exercise parameters (aerobic capacity on cardiopulmonary exercise testing expressed as peak VO_2_ and distance in metres walked in 6 minutes). The Kansas City Cardiomyopathy Questionnaire 12 item version (KCCQ-12) was the only survey tool used and was reported in a single study as a measure of patient reported outcomes.^13^

The duration of follow-up after SGLT2i therapy ranged from a 12-week observation period to a median follow-up duration of 19 months (range 14-31 months). Pooled analysis was restricted to studies of similar design (cohort studies examining pre/post intervention effect)(n=8). The frequency of outcome metrics reported across studies is shown in Figure 2. The most frequently reported outcome metric was NT-proBNP (n=6 studies, 60%) followed by GLS (n=5 studies, 50%), BNP and FAC (n=4 studies, 40%, respectively).

**Figure 2.**
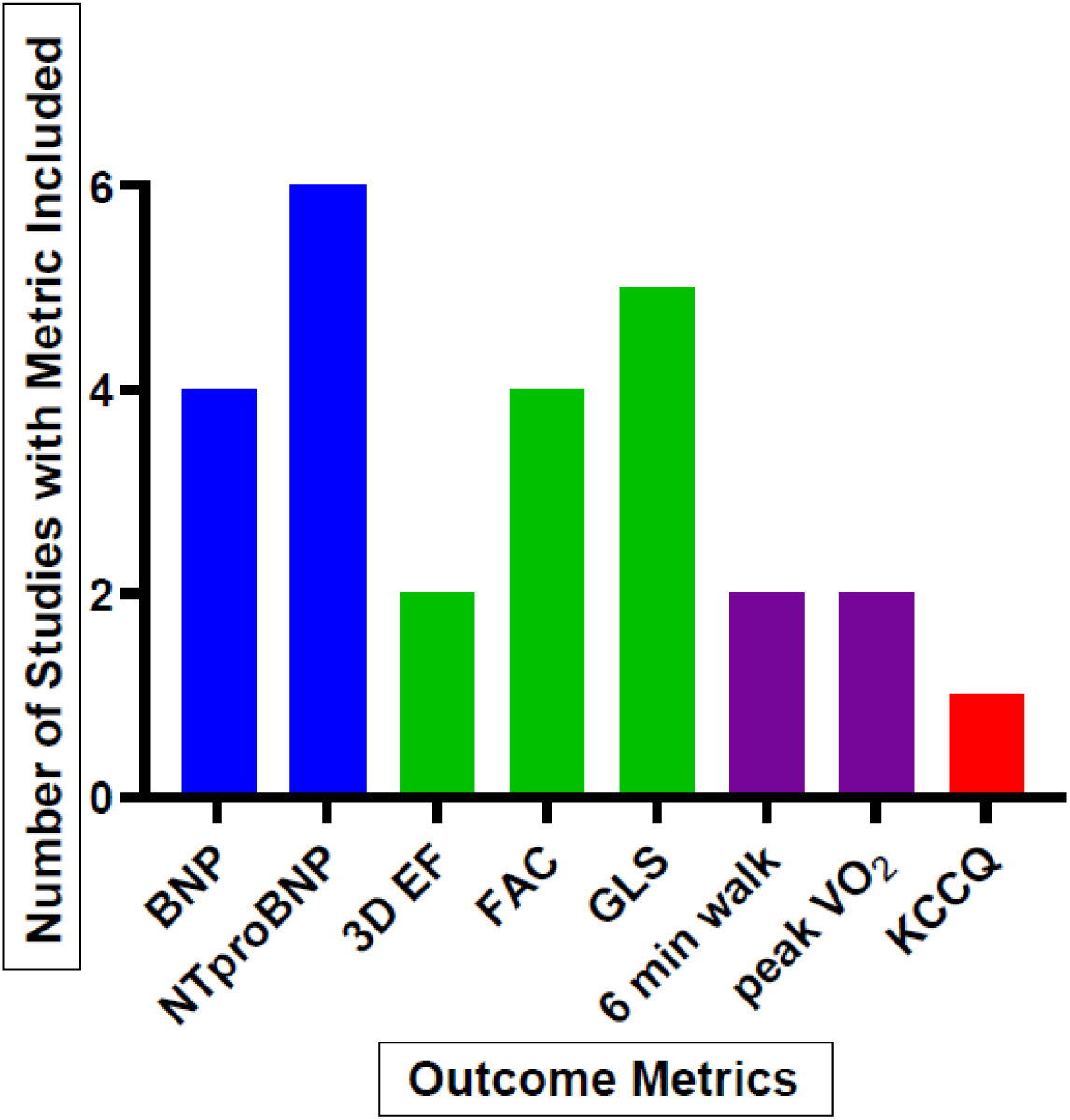
Number of Studies Reporting Outcome Metrics. BNP, brain natriuretic peptides; EF, ejection fraction; FAC, fractional area change; GLS, global longitudinal strain; KCCQ, Kansas City Cardiomyoathy Questionnaire; NT-proBNP, n-terminal pro brain natriruretic peptide; RV, right ventricle; peak VO_2_, peak aerobic capacity; 3D, three dimensional

Forest plots for the effects of SGLT2i therapy on serum NT-proBNP and BNP levels are displayed in Figure 3. The heterogeneity for the pooled effect of SGLT2i therapy was moderate for NT-proBNP (n=5 studies; I^2^=47%) and high for BNP (n=3 studies; I^2^= 90%). The random effects model estimated mean differences in NT-proBNP and BNP that favored SGLT2i therapy but which were not statistically significant (mean difference -240 pg/mL [95% CI -516, 45] and -52 pg/mL [95% CI -129, 26], respectively)(Figure 3). With respect to the 5 individual study results for NT-proBNP, a statistically significant change in NT-proBNP was reported following SGLT2i therapy in 3 studies (Table 1)^7,8,15^ and a non-statistically significant change favoring SGTL2i therapy was observed in one study.^10^ In contrast, there were no statistically significant changes in any of the studies assessing BNP. Of note, these studies had the smallest sample sizes and additionally included patients who were well-optimized on medical therapy with a normal BNP level prior to SGLT2i therapy and were therefore unlikely to have an observed impact of SGLT2i therapy.^10,12,14,16^

**Figure 3.**
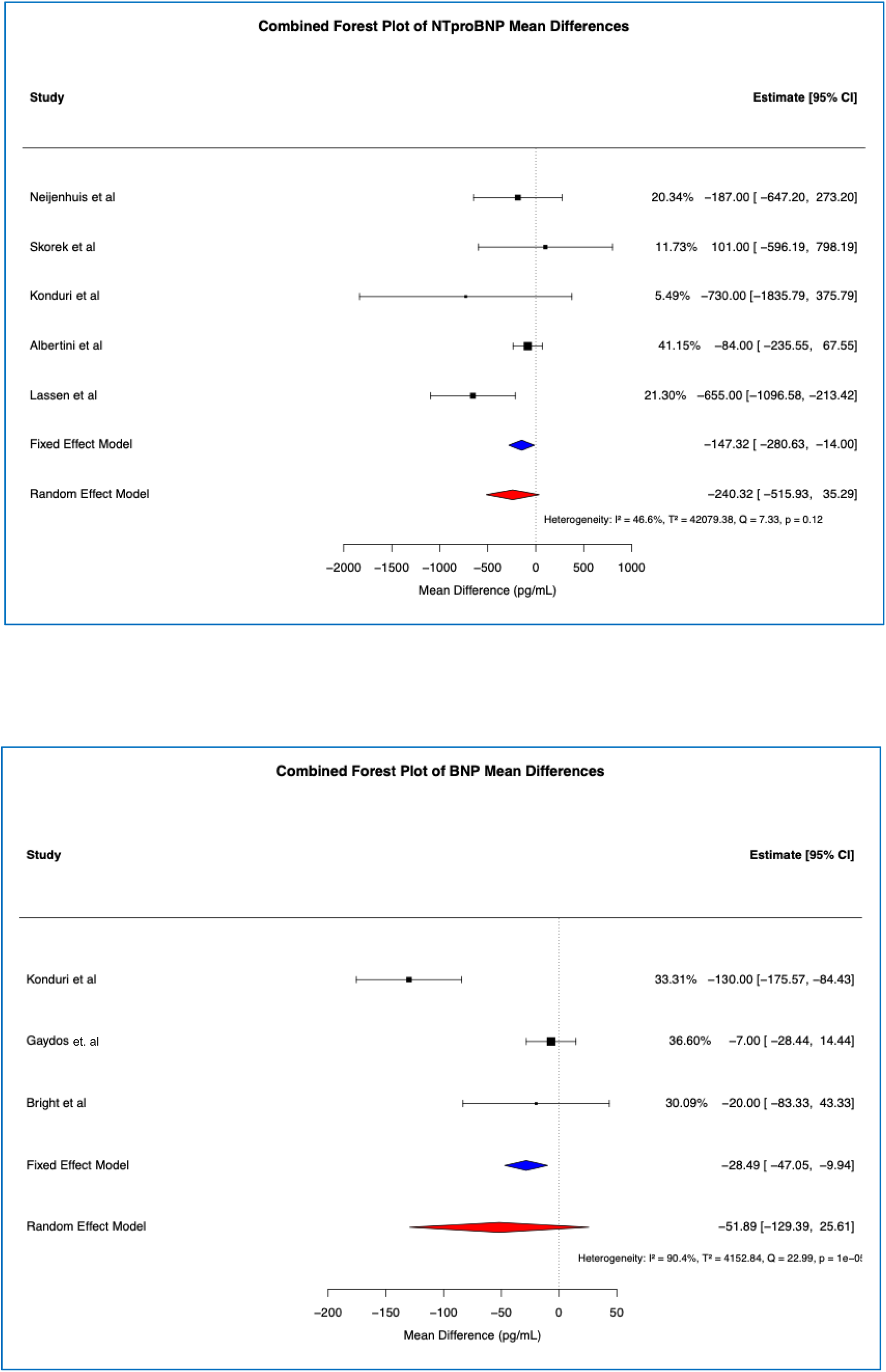
Forest plot for NT-proBNP (top panel) and BNP (bottom panel)

The effects of SGLT2i therapy on GLS and FAC on Forest plots are shown in Figure 4. The heterogeneity for measures of systolic function following SGLT2i therapy on GLS (n=4 studies) and FAC (n=3 studies) were low, I^2^ 0% for both. There was a statistically significant difference in GLS following SGLT2i treatment (mean difference -1.6 [95% CI -2.4, -0.9]), consistent with an improvement of ventricular systolic function in the longitudinal direction as measured by strain. There was no apparent improvement in FAC following SGLT2i treatment (mean difference +1.86 [95% CI -6.2, 9.9]) (Figure 4). Further review revealed a statistically significant change in both GLS and FAC in 2 studies,^7,13^ a non-statistically significant numerical change in GLS and FAC in 1 study, and a non-statistically significant change in GLS in 1 study (Table 1).^11,12^

**Figure 4.**
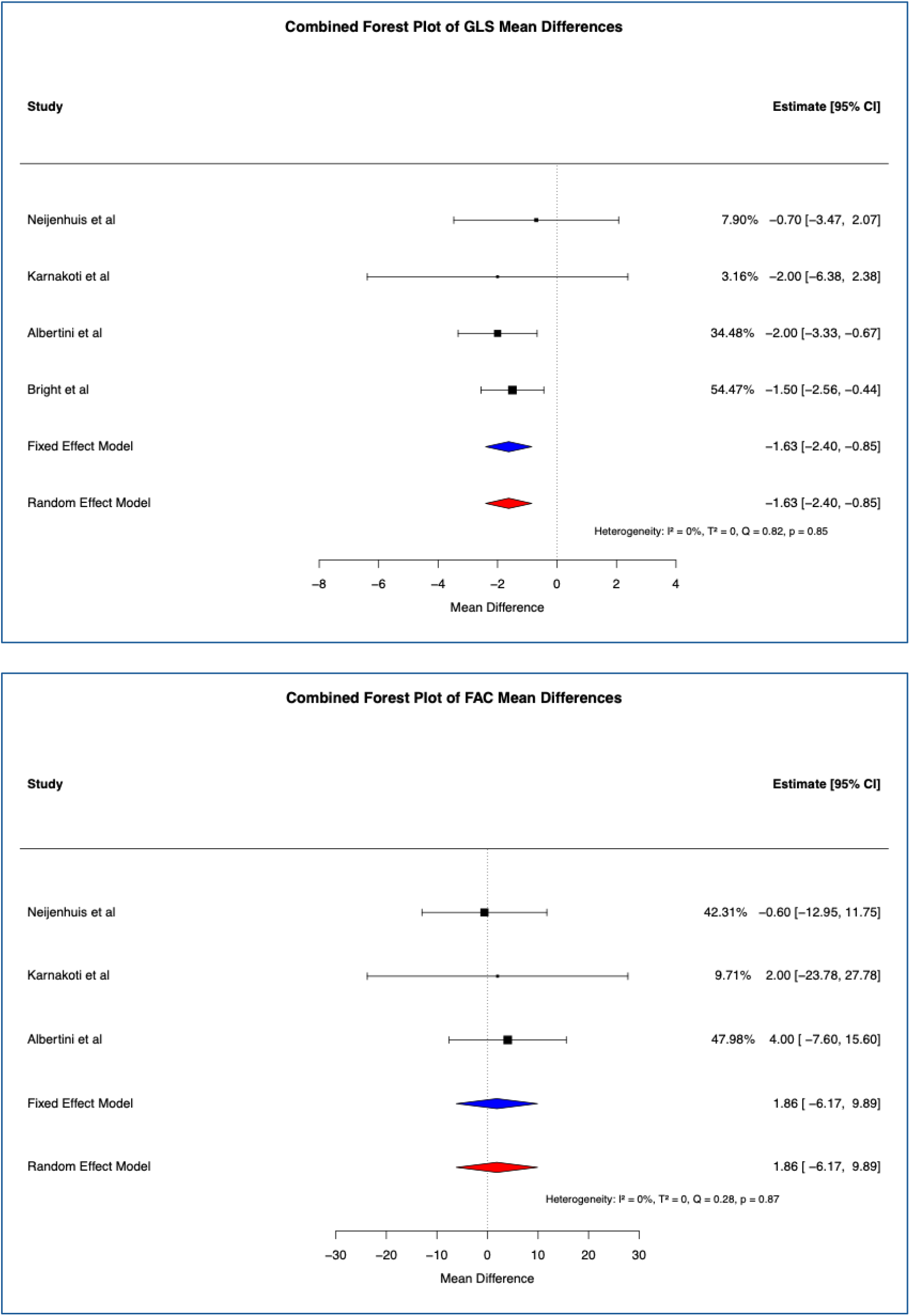
Forest plot of GLS (top panel) and FAC (bottom panel)

Although peak VO_2_ appeared to have a statistically significant increase post SGLT2i therapy, this was based on a subgroup group of patients with relatively small numbers along with large amounts of missing data.^9^ Similarly, another study with small numbers and a large amount of missing data found an increase in peak VO_2_ which was not statistically significant (Table 1). The 6 minute walk test was used as an outcome measure in 2 studies; there was a statistically significant increase in distance covered post SGLT2i therapy in one study and a non-statistically significant increase in another.^7,13^

Finally, the KCCQ survey results, reflecting patient reported outcome measures, were only reported in one study. This study demonstrated a statistically significant improvement in KCCQ-12 scores following SGLT2i therapy.^13^

### Drug-related serious adverse events

With the exception of the study by Bright et al (which reported on GDMT for a variety of agents in addition to SGLT2i),^12^ all studies recorded occurrences of adverse events, if any, related to administration of SGLT2i. There were no serious adverse events related to SGLT2i therapy. In the study by Neijenhuis et al, 1 patient had a urinary tract infection which was treated with antibiotics.^8^ In the study by Konduri et al one patient was noted to have transient acute kidney injury related to diuresis.^10^ In the study by Gaydos et al a single patient had pre-existing joint and abdominal pains which were felt to be unrelated to SGLT2i therapy but were the reasons listed for discontinuation of SGLT2i therapy.^14^ In the remaining studies there were no SGLT2i-related adverse events or safety concerns reported.

### Risk of bias

Risk of bias for cohort studies was assessed using the Cochrane ROBINS-I tool which revealed that half of the studies were at serious risk and the remaining were at moderate risk of bias (Supplementary Figures 2 and 3). The main issues pertained to patient selection bias, potential for confounding (i.e. lack of adjustment for severity of HF at study entry, interaction between SGLT2i and other HF medications), substantial amounts of missing data and questionable outcome ascertainment (i.e. unblinded review of imaging studies for assessment of ventricular function). The risk of bias for the one clinical trial in this analysis was assessed using the ROB-2 tool suggesting a high risk of bias (of note the trial was published as a short research letter which imposed limitations on extent of methodologic description). The Newcastle Ottawa Score results are shown as a complementary assessment of bias in cohort studies (Supplementary Figure 4). Based on review and analysis of the funnel plots, no risk of publication bias for the NT-proBNP, BNP, GLS and FAC outcomes measures (Supplementary Figure 5); specifically, all Egger test values for the outcomes measured were found to be non-significant.

## DISCUSSION

Given the increased morbidity and mortality related to HF in complex ACHD, identification of effective medical therapies to support and prevent ventricular failure are critically important. However, these have yet to be elucidated specifically in the setting of sRV or SV anatomies. Medical therapies that have proven to be successful in the left ventricle (LV) of a biventricular circulation, such as angiotensin-converting-enzyme inhibitors, mineralocorticoid receptor antagonists, and beta-blockers, have yielded disappointing results in the sRV and SV populations, possibly a reflection of the distinct functional and morphological features inherent in each of these complex anatomies.^21,22^ Because medical strategies cannot be extrapolated from the failing LV in a biventricular circulation to the failing RV or the SV, robust studies are specifically needed in these vulnerable populations. Emerging data suggest that potential benefits of SGLT2i in ACHD are plausible, but existing studies are few, are often prohibitively small, and tend to include mixed populations without clear stratification by anatomic subtype.

In this systematic review and meta-analysis of SGLT2i therapy in patients with complex CHD defined as sRV or SV anatomies, there appeared to be an improvement in GLS following SGLT2i treatment in some studies although observations were limited by missing datapoints in many studies and the small numbers of patients studied overall. Nevertheless, the magnitude of absolute GLS change noted in our study (mean difference 1.6%) was similar to a recent meta-analysis evaluating change in cardiac function parameters in a normal biventricular circulation (mean difference 1.17%).^23^ Furthermore, there were only scant data on changes in 3D EF on echocardiography and no studies reporting CMR imaging measurements. There did not appear to be a significant improvement in FAC and there were no significant differences in either NT-proBNP or BNP serum levels following SGTL2i therapy. There were insufficient data to draw conclusions regarding impact of SGLT2i therapy on adverse clinical outcomes, change in exercise parameters and/or patient reported outcomes.

### SGLT2i and acquired HF

The SGLT2i class of medications were initially developed for management of diabetes, but have since resulted in a radical transformation in the management of HF. These agents are now endorsed globally as Class I guideline-directed medical therapy for HF across a range of ejection fractions in adults with a biventricular circulation, with or without diabetes. ^2,24^ These recommendations arise from compelling evidence from robustly conducted clinical trials demonstrating that SGLT2i therapy is effective for reducing cardiovascular death and HF hospitalizations.^25–29^ In fact, the totality of evidence from 5 large SGLT2i randomized controlled trials with >22,000 HF participants demonstrated that SGLT2i reduced the risk of cardiovascular death or HF hospitalization by 23%, cardiovascular death by 13%, and all-cause mortality by 8%.^30,31^ Of note, the benefits of SGLT2i on clinically relevant outcome measures in adults with HF are remarkably consistent across sub-groups of age, sex, background pharmacotherapy, diabetes status, and baseline measures of HF including HFpEF and HFrEF (such as B-type natriuretic peptide [BNP] levels). Notably, individuals with ACHD patients were not included in any of these landmark HF trials.

### Treatment of HF in the ACHD population

Although the most recent guidelines for HF management published by the American Heart Association/American College of Cardiology and the European Society of Cardiology strongly support the use of SGLT2i in patients with LV dysfunction in the context of structurally normal hearts,^5,6^ these guidelines do not address HF treatment in ACHD. It should be noted that in the ACHD population there is a broader array of pathophysiologies which may contribute to HF, some of which include maladaptive response of the sRV or SV to systemic pressures, post-operative changes which can induce constriction or restriction, and/or mechano-electrical disease which may result in ventricular dyssynchrony.

Although patients with complex ACHD are at substantially elevated risk of HF with advancing age, the totality of evidence supporting targeted medical therapies is scant. One systematic review of conventional HF therapies in adults with a sRV (focused on the clinical impact of angiotensin-converting enzyme inhibitors, angiotensin-receptor blockers, beta-blockers and aldosterone antagonists) did not find evidence of benefit on cardiovascular imaging or exercise testing (SGLT2i not included in this study). Of note, the methodologic quality of the majority of studies in that review was felt to be suboptimal given low power, incomplete follow-up and deficiencies in study design.^22^ Even less is known about the role of this newer class of HF medicines, SGLT2i, in ACHD-HF, both in terms of efficacy and safety. Only a single systematic review has been published which included 8 studies with a total of 287 patients and examined SGLT2i use across all forms of ACHD (inclusive of mild, moderate and severe complexity disease).^32^ However, no systematic reviews have specifically focused on complex ACHD patients with complex (sRV or SV) anatomies. As such, the potential role of SGLT2i in managing HF in these high-risk ACHD subgroups remains poorly defined.

### Strengths and weaknesses of studies selected for analysis

There are several salient strengths of the studies which were selected for inclusion in this systematic review. Firstly, there was a relative homogeneity in anatomies as all the studies focused on, or at least contained patient level data pertaining to, adults treated with SGTL2i therapy with either sRV or SV forms of ACHD. Secondly, the studies entered in the analysis were all contemporary studies published between 2022 and 2025, reflecting the relative novelty of this class of medication in the treatment of ACHD-HF and rendering the study results generalizable to the current era of patient care. There was reasonably complete follow-up, both in terms of patient numbers and duration of observation time, in all the studies included in this systematic review. Finally, biochemical and imaging outcomes are objective measurements with extensive clinical validation.

However, there are various weaknesses in the studies selected which are worthy of mention. These include issues pertaining to study design and potential for bias. Of the ten studies, there was only 1 randomized controlled trial;^7^ this was an open label study with associated risks of bias and was reported as a short research letter which limited the amount of detail which could be included (ie method of randomization is not provided, deviations from protocol not described etc). The majority of studies, 80% in total, were cohort studies (5 retrospective and 3 prospective) describing changes pre-versus post-SGLT2i therapy, and only these were entered into the meta-analysis. There was one case series which reported on a convenience sample of 5 patients and could not be appropriately assessed for bias.^16^ Overall published cohorts were relatively small in size (the largest study had only 32 patients). Furthermore, if studies reported on a range of ACHD anatomies or included children along with adults, then only the subset of patients who were adults with sRV or SV lesions were included in the analysis, which further limited study power (40% of the studies had only 5-10 patients who met inclusion criteria). Of note, limited study power precluded meaningful subgroup analysis and therefore heterogeneity demonstrated in the meta-analysis could not be further explored.

Major adverse cardiovascular events were limited to report of HF hospitalizations in 2 studies and only one study included patients reported outcomes in the form of the KCCQ-12 questionnaire.^13^ Therefore, surrogate outcome measures (serum and imaging biomarkers) were included into the meta-analysis instead. While the NT-proBNP and BNP serum measures are biochemical assays which detect HF, these assays differ and results therefore cannot be directly compared. It is notable that SGLT2i therapy appeared to have an impact on NT-proBNP levels but not BNP, for reasons which are not clearly apparent, but may relate to differences in population characteristics and/or sample size (larger in NT-proBNP and smaller for BNP studies). Most studies did not account for severity of HF at the time of initiation of SGLT2i treatment (if biochemical markers of HF are normal at the time of initiation of therapy as seen in several studies,^12,14,16^ then it is not surprising that the impact of medical therapy for HF as measured by this biochemical marker would be negligible). Although CMR is considered the most robust imaging modality for assessment of ventricular systolic function (and the reference standard for measurement of EF), the 5 studies which described imaging outcomes reported echocardiographic measures only. In the absence of CMR assessment of systemic ventricular EF (which would provide the highest fidelity measurement), only 2 studies reported on 3D EF using echocardiography and the remaining studies reported relatively less robust 2D of ventricular function measured in a single plane, namely FAC and GLS.

There was further heterogeneity with respect to type of medications used and the severity of HF being treated. While 4 studies evaluated the impact of dapagliflozin only, the remaining 6 studies evaluated the impact of either dapagliflozin or empagliflozin without adjustment for the type of SGLT2i used (to address the possibility that there may be a differential effect of dapagliflozin versus empagliflozin). Additionally, patients were on various combinations of added HF medications across studies which could either potentiate or attenuate the impact SGLT2i, and this was not adequately adjusted for in any of the reports. Adherence to the prescribed SGLT2i therapy was only ascertained in one of the studies (by telephone calls to study participants).^12^ Furthermore, although the range of follow-up durations was felt to be adequate overall, there is a possibility that a shorter duration of therapy (i.e. 12 weeks) may in fact be too short to fully appreciate the impact of SGLT2i treatment.

Finally, the Cochrane risk of bias tools demonstrated that there was important risk of bias to be considered (moderate in 4 of the cohort studies, high in 4 of the cohort studies and high in the single trial). Specifically, external validity may be limited as all studies occurred in tertiary care settings in high income countries (Table 1, Supplementary Figure 1). Internal validity may be limited as the majority of studies did not include blinding during reporting of imaging or exercise testing. Survival bias may affect the studies with a pre/post intervention design as only patients who presented for follow-up post drug intervention are reported on. Selection bias may affect results if SGLT2i therapy was only offered to a proportion of patients in the eligible population (perhaps these are sicker patients, patients able to afford the drug, or are patients most likely to come for follow-up). Misclassification bias may occur if a patient is thought to have received SGLT2i but, in fact, was under-dosed or inconsistently treated such that a therapeutic effect would be unlikely. Confounding factors were not adjusted for in any of the included studies.

### Comparison with other published studies

To date, the only systematic review on the topic of SGLT2i for treatment of chronic HF in ACHD was published by Das et al in 2025 which reported on a broad spectrum of ACHD anatomies.^32^ The present study furthers the published literature in several ways. In this current study a more stringent approach is applied to the inclusion criteria, focusing only on systemic ventricles with “non-LV type” morphologies. This distinction is of importance as previously published HF literature suggests that medical therapies have a differential response in the LV versus non-LV morphologies (such as sRV and SV lesions). Although it would be preferable to assess the differential effects of SGT2i therapy on the sRV independently from the SV, at the present time there are insufficient patient numbers for this level of stratification.

In the construct of this systematic review the decision was made to focus on peer-reviewed publications excluding abstracts. With this approach the aim was to incorporate and review only the highest quality data although it should be acknowledged that this may have imposed an element of publication bias as abstracts and the grey literature were excluded (which could represent exclusion of relevant non-published negative studies). Although inclusion of published abstracts would have resulted in a larger number of patients studied, this approach could potentially compromise the overall calibre of data and these were ultimately excluded. Only English language publications were included, which led to exclusion of 7 non-English language publications; following review of the abstracts of these publications it was determined that these would be unlikely to have substantial impact on the systematic review as a whole. There appears to be a low risk of publication bias based on the funnel plots with the corresponding Egger test, which may be reassuring or may reflect the small number of studies included.

### Strengths and weakness of the current meta-analysis

The low heterogeneity between studies with measures of systolic function suggests that the observed differences are unlikely to be due to differing patient populations or differences in study design but may also be attributed to the small number of available studies included in the meta-analysis. The change in GLS, consistent with improvement in ventricular function after SGLT2i therapy was statistically significant, pointing towards the fact that medical therapy may contribute to a measurable improvement in heart function; the clinical relevance of this observation remains uncertain. In contrast, there were no clinically significant improvements in FAC following SGLT2i therapy and it is uncertain if this was due to small number of studies with modest number of patients or technical differences in measurement (of note, FAC can be a more challenging imaging measurement to make as compared with GLS).

There was substantial heterogeneity between studies measuring the pooled effect of NT-proBNP and BNP following SGLT2i therapy and mean differences based on random effect models were not found to be statistically significant. It is important to note that at least half of the variability noted in these measures could be attributed to between study heterogeneity. Based on the published literature to date, it is widely accepted that serum biomarkers such as NT-proBNP and BNP are relatively robust endpoints and although not statistically significant in this suggest these assays are likely worthy of further study in larger studies with a randomized controlled design.

The results of the meta-analysis may be limited by several factors. Small sample sizes were made smaller still in cases where patient level data was extracted to meet the pre-specified inclusion criteria (i.e. only data from adults were included, and pediatric data were excluded). Although each is considered a form of complex CHD, sRV and SV reflect differing ventricular anatomies and different physiologic states. In many cases the effect estimates were not reported with a measure of precision or statement of statistical significance, which may be attributed to under-powered studies with a relatively high number of missing data-points. Variability in effect measures was also noted - the only outcome measure which was reported in at least 5 studies was NT-proBNP; BNP was only reported in 3 studies but could not be directly compared to NT-proBNP due to differences in assay characteristics. Cardiac imaging outcomes were similarly limited, with GLS reported as an endpoint in 4 studies and FAC in 3 studies (Figure 2). Heterogeneity between biochemical serum assays was apparent but could not be further assessed with subgroup analyses due to limited patient numbers. Finally, potential for bias in the meta-analysis is substantial as demonstrated by the formal Cochrane risk of bias evaluation; more than half of the studies were high risk of bias with the remainder were at moderate risk.

Worthy of mention is one study in this systematic review (not entered into the meta-analysis of pre/post mean difference studies) which was an open-label randomized trial which observed a mean difference in NT-proBNP in the treatment group of -33 (-87, -1) compared with 5 (0, 15) in the control group which was of borderline statistical significance (p=0.05). In the same trial there was a statistically improvement in GLS in the treatment group of -1.6 (-3.3, -0.7) compared to the control group of -0.1 (-0.2, 0.4)(p=0.0001).^7^ These results are supportive of the results observed in our meta-analysis.

Overall, data for inclusion in the meta-analysis were limited. Outcomes were surrogate markers of HF (serum biomarkers and imaging measures of function) rather than truly adverse cardiovascular outcomes. Individual participant level data collection could be considered as a future direction of investigation but was beyond the scope of the present study. The results of the meta-analysis point towards the need for larger studies with a randomized controlled design for further assessment of the impact of SGT2i therapy on HF management in complex ACHD.

### Implications for clinical practice

The results of this study may have implications for clinical practice. For those providing care at the individual level, an important message to emerge is the absence of any reportable significant adverse events as a result of SGLT2i therapy. Therefore, even without clear evidence of benefit, SGLT2i therapy is unlikely to do harm in patients with HF in the context of sRV or SV lesions. Furthermore, there is a signal in the data suggesting select imaging measures of RV function may improve following SGLT2i therapy although further study is required to explore the clinical significance of this observation.

Of note, at least 12 million people are living with CHD world-wide and most deaths have been observed in low and low-middle sociodemographic index quintiles.^33,34^ An important observation from this study is the striking absence of studies from any middle-income or low-income countries. In this regard, drug cost and availability should be critically evaluated in addition to support for research infrastructure in middle and low-income countries. Additionally, SGTL2i availability appears to be linked with tertiary care settings, and it would be worthwhile exploring whether greater availability in community-based healthcare practices would be feasible, practical and/or relevant. Adequate drug availability across all sociodemographic sectors should be a goal of future therapy.

### Directions for future research

There is currently an absence of data pertaining to the impact of SGLT2i therapy on major adverse cardiovascular events in adults with non-LV morphology CHD, namely cardiovascular-related mortality, cardiac transplantation and hospital admission for HF. Additionally, there is a relative paucity of data pertaining to the impact of SGT2i therapy on patient reported outcomes. Finally, there is an absence of data comparing the relative effect of SGLT2i therapy according to ventricular subtype, specifically LV versus RV versus SV.

Further research should aim for more robust cardiovascular imaging measurements. Ideally all measurements on imaging studies would occur in a blinded fashion according to a predefined study protocol. Non-LV-type ventricular morphologies should have functional measurements derived from CMR rather than echocardiography, as the former is considered the reference standard for non-invasive assessment of cardiac function.

The many weaknesses of the ten selected studies outlined above underscore the need for well-designed future studies - ideally larger, double-blinded, randomised controlled trials across multiple centres with attention to internal and external validity of results. Generalizability of results requires attention to recruitment of patients across the spectrum of health-care settings, specifically low-, middle-and high-income countries.

## CONCLUSIONS

In this systematic review and meta-analysis of SGLT2i therapy in complex ACHD, there appeared to be an improvement in parameters of systemic ventricular systolic function but not serum biomarkers after SGLT2i treatment. There were insufficient data to draw conclusions regarding change in exercise parameters, patient reported outcomes or major adverse cardiovascular events. Notably there were no significant drug-related adverse events reported in any of the ten studies included in this review. All studies were performed in tertiary care centres in high income countries. Future studies would ideally be larger, double-blinded, randomized controlled trials designed to maximize external and internal validity of results across a variety of healthcare settings and geographical locations.

## Data Availability

Data will be made available upon reasonable request

## ABBREVIATIONS

ACHD: adult congenital heart disease
BNP: brain natriuretic peptide
CHD: congenital heart disease
CMR: cardiovascular magnetic resonance imaging
EF: ejection fraction
FAC: fractional area change
GDMT: guideline directed medical therapy
GLS: global longitudinal strain
HF: heart failure
NT-proBNP: N-terminal pro brain natriuretic peptide
LV: left ventricle
RV: right ventricle
sRV: systemic right ventricle
SGLT2i: sodium glucose co-transport inhibitor
SV: single ventricle

## SUPPLEMENTARY DATA

### APPENDIX: Details of the search strategies employed

#### Search Strategy 1: Pubmed, Embase, SCOPUS, and Cochrane Databases

Keywords:

1. “Sodium-glucose cotransporter 2 inhibitor” or “SGLT2” or “SGLT2i” or Empagliflozin or Dapagliflozin or Canagliflozin or “guideline directed medical therapy” or “heart failure therapy” or “flozin” AND
2. “Fontan” or “systemic right ventricle” or “Fontan circulatory failure” or “univentricular heart” or “single ventricle” or “Mustard surgery” or “Senning” or “congenitally corrected transposition of the great arteries” or “atrial switch” AND
3. “Symptoms” or “New York Heart Association” or “B-type natriuretic peptide” or “BNP” or “exercise test” or “6 minute walk” or “hospitalization” or “systolic function” or “ejection fraction”

MESH headings:

“Sodium-Glucose Transporter 2 Inhibitors” heading; entry terms SGLT-2 Inhibitors, SGLT 2 Inhibitors, SGLT-2 Inhibitor, Inhibitor, SGLT-2, SGLT 2 Inhibitor, Sodium-Glucose Transporter 2 Inhibitor, Sodium Glucose Transporter 2 Inhibitor, SGLT2 Inhibitor, Inhibitor SGLT2, Gliflozins, Gliflozin, SGLT2 Inhibitors

AND

“Fontan Procedure” heading; entry terms procedure Fontan, Fontan operation, operation Fontan, Fontan palliation, palliation Fontan, Fontan circulation, circulation, Fontan, Fontan circuit, circuit Fontan

“Systemic“ [All Fields] AND (“heart ventricles“[MeSH Terms] OR right ventricle [Text Word])

#### Search Strategy 2: Medline

1. exp Sodium-Glucose Transporter 2 Inhibitors
2. sodium glucose transporter 2 inhibitor*.tw.
3. sodium-glucose cotransporter 2 inhibitor*.tw.
4. sodium-glucose co-transporter 2 inhibitor*.tw.
5. sglt2*.tw.
6. sglt-2*.tw.
7. dapagliflozin.tw.
8. empagliflozin.tw.
9. canagliflozin.tw.
10. 1 or 2 or 3 or 4 or 5 or 6 or 7 or 8 or 9 (17340)
11. heart defects, congenital/ or “transposition of great vessels”/ or “congenitally corrected transposition of the great arteries”/ or double outlet right ventricle/ or univentricular heart/
12. Fontan Procedure
13. Ventricular Dysfunction, Right
14. fontan*.tw.
15. (congenital adj3 (heart or cardiac) adj3 (disease* or defect* or malform* or abnormalit*)).tw.
16. (right adj3 (ventricular or ventricle*) adj3 (fail* or dysfunction)).tw.
17. CHD.tw.
18. ACHD.tw.
19. 11 or 12 or 13 or 14 or 15 or 16 or 17 or 18
20. 10 and 19

**Supplementary Figure 1.**
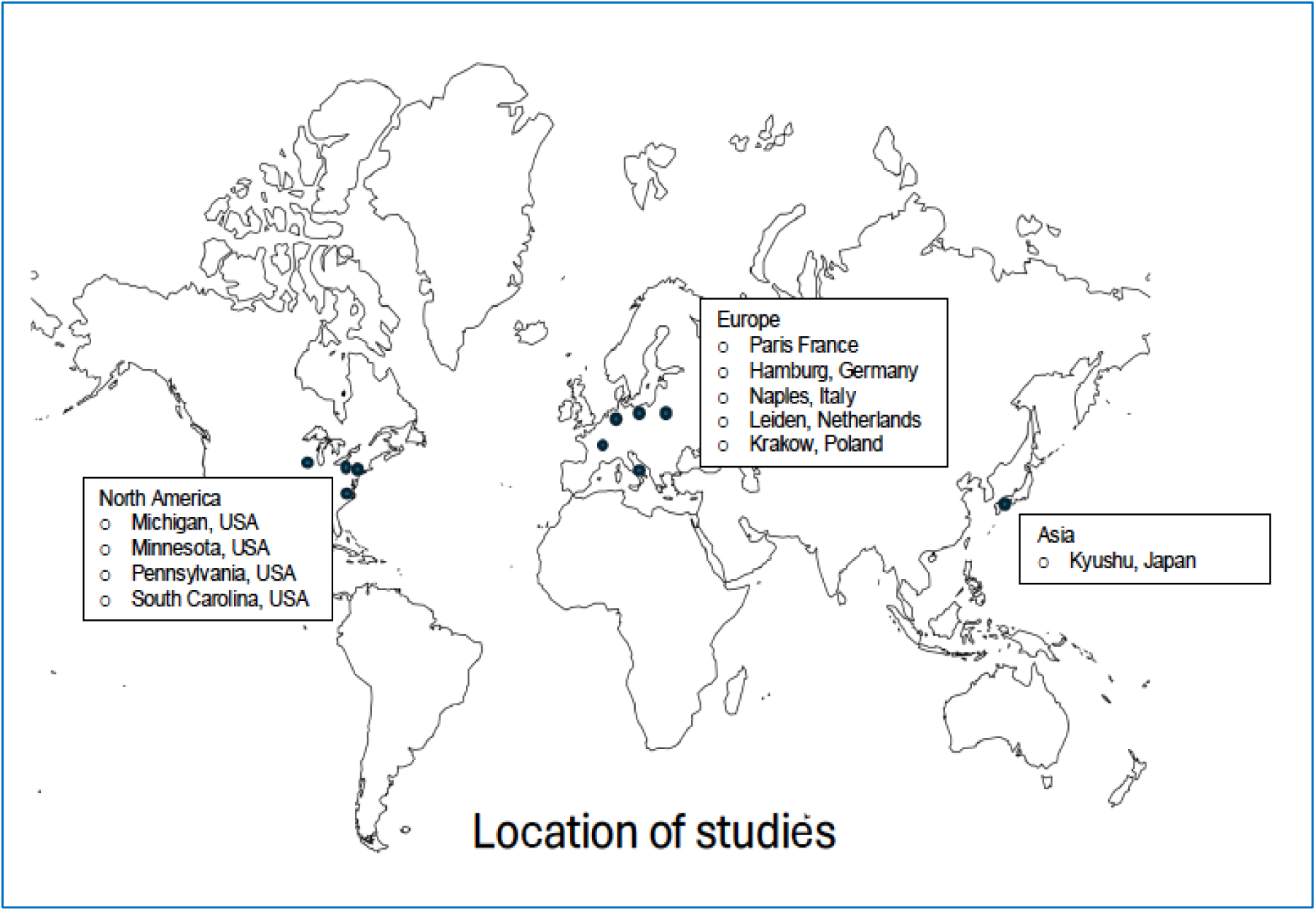
Location of Studies Selected for Inclusion.

**Supplementary Figure 2.**
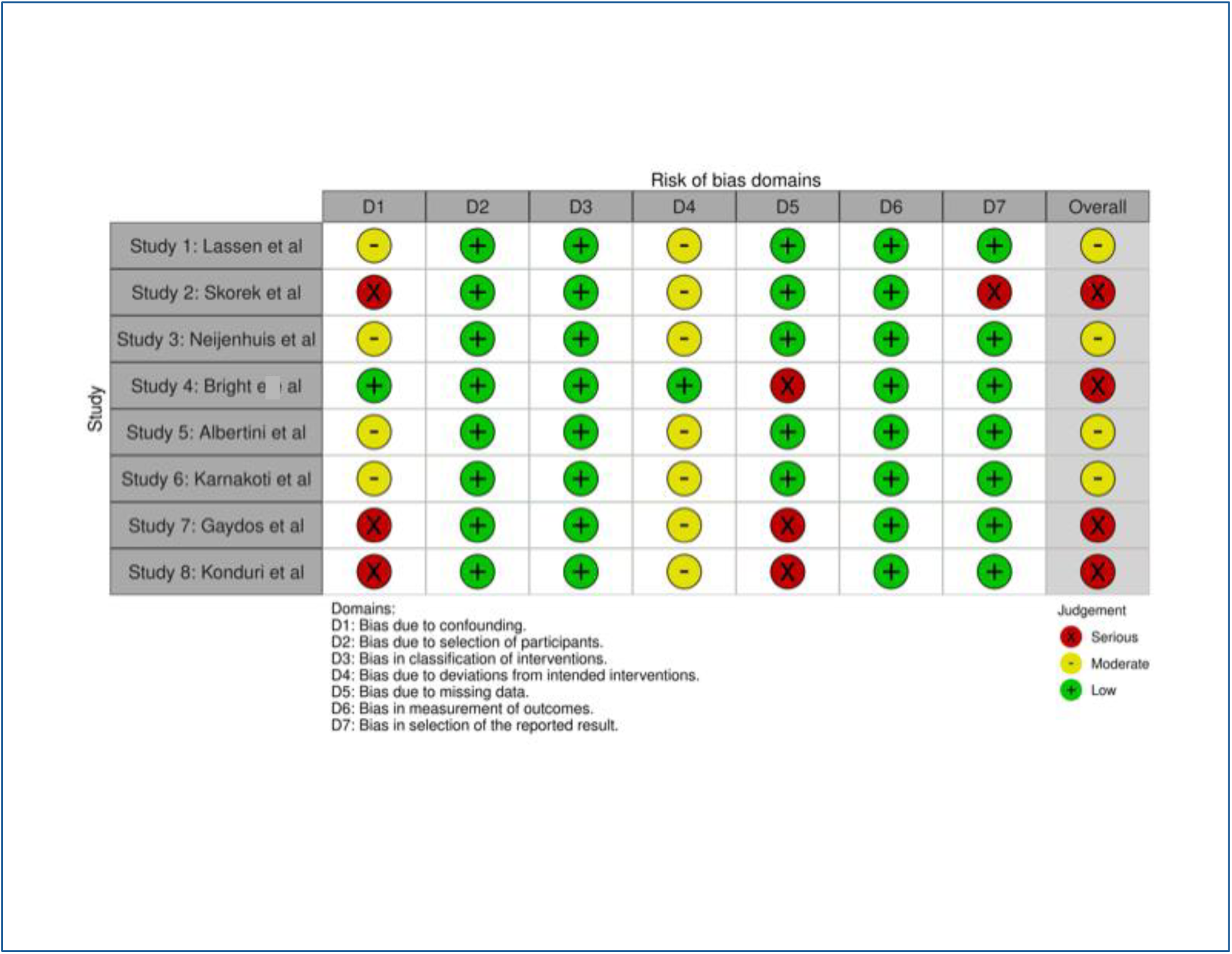
ROBINS-I risk of bias tool for cohort studies. For the ROBINS-I tool, 7 domains were reviewed for each study: (1) risk of bias due to confounding (2) risk of bias in classification of interventions (3) risk of bias in selection of participants into the study (4) risk of bias due to deviations from intended interventions (5) risk of bias due to missing data (6) risk of bias from measurement of the outcome (7) risk of bias in selection of the reported result. Each category received an assignment of either “yes”, “partially yes”, “no”, “partially no” or “insufficient data” and the study was ultimately assigned a grade of overall risk of bias “low”, “moderate”, “serious” or “critical”. Legend: Risk of bias low (green), medium (yellow), high (red)

**Supplementary Figure 3.**
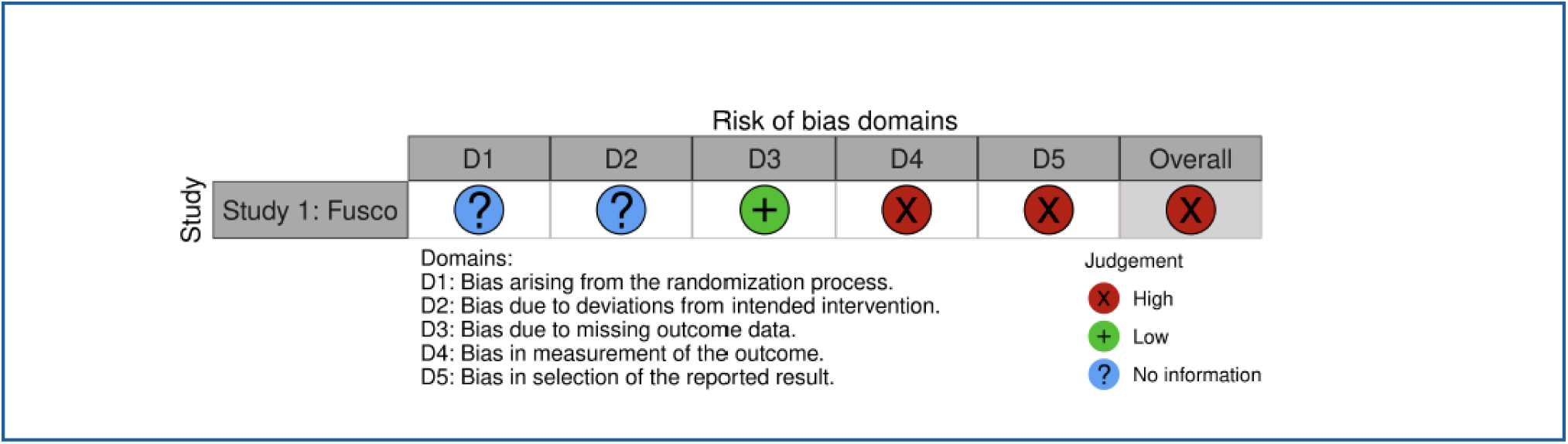
ROB-2 risk of bias tool for randomized studies. For the ROB-2 tool for assessment of interventional studies, the following 5 domains were evaluated: (1) bias related to the randomization process (2) bias related to deviation from the intended intervention (3) bias related to missing outcome data (4) bias related to measurement of the outcome and (5) bias related to reporting outcomes. In addition, a global summary score was assigned for the trial. Each category was designed as “low” risk, “some concern”, high” risk or “no information”. Legend: Risk of bias low (green), medium (yellow), high (red)

**Supplementary Figure 4.**
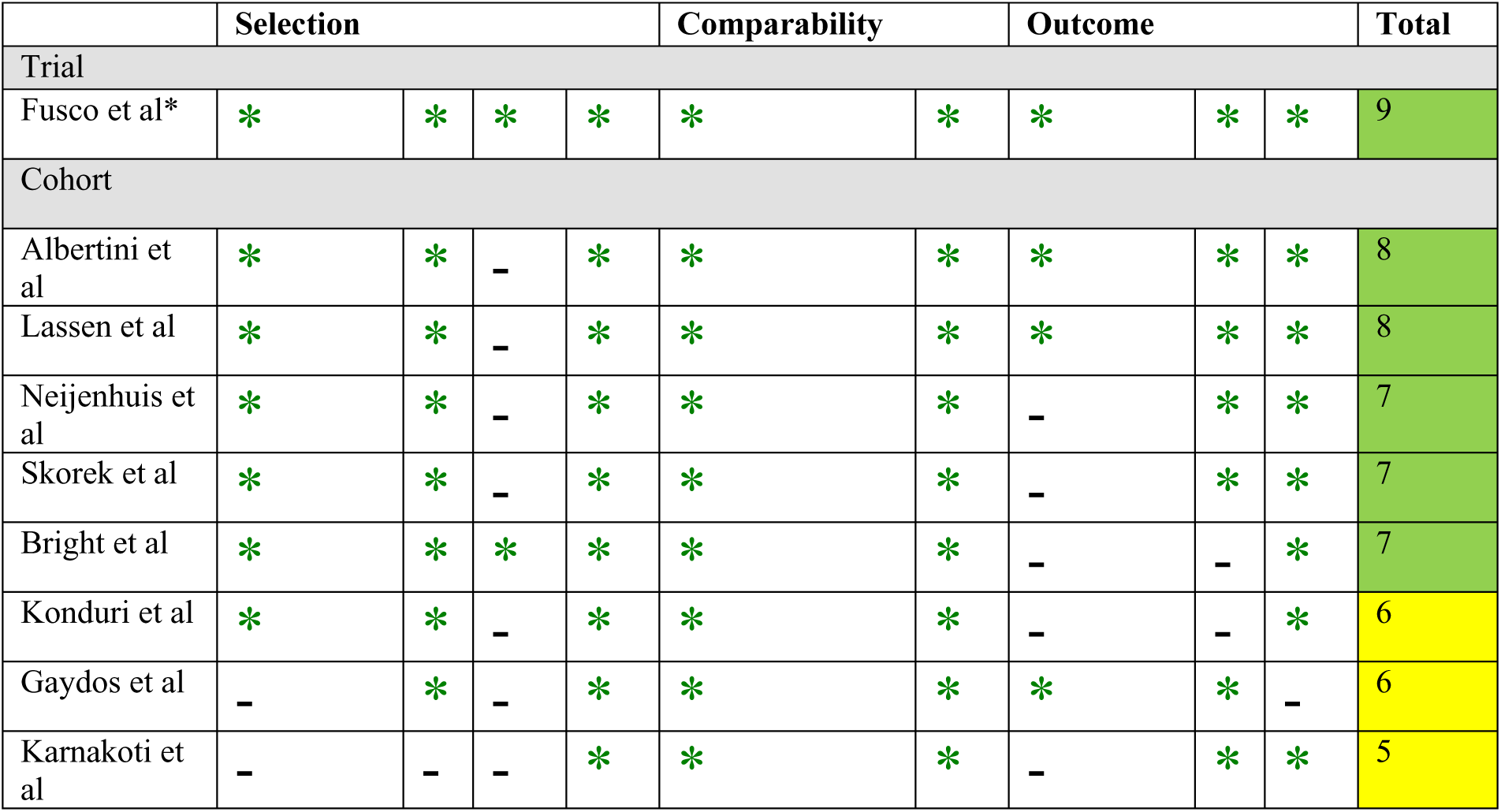
Newcastle Ottawa Scale for risk of bias assessment in cohort studies. At total of 9 specific domains were reviewed encompassing (1) selection (2) comparability and (3) outcomes. Each study received a score from 0 to 9 and bias was accordingly assigned (7-9 low risk, 4-6 medium risk and 1-3 high risk of bias). Legend: Risk of bias low (green), medium (yellow), high (red) *although included here, Fusco is a trial – and will have additional review of risk of bias based on the Cochrane criteria in the text

**Supplementary Figure 5.**
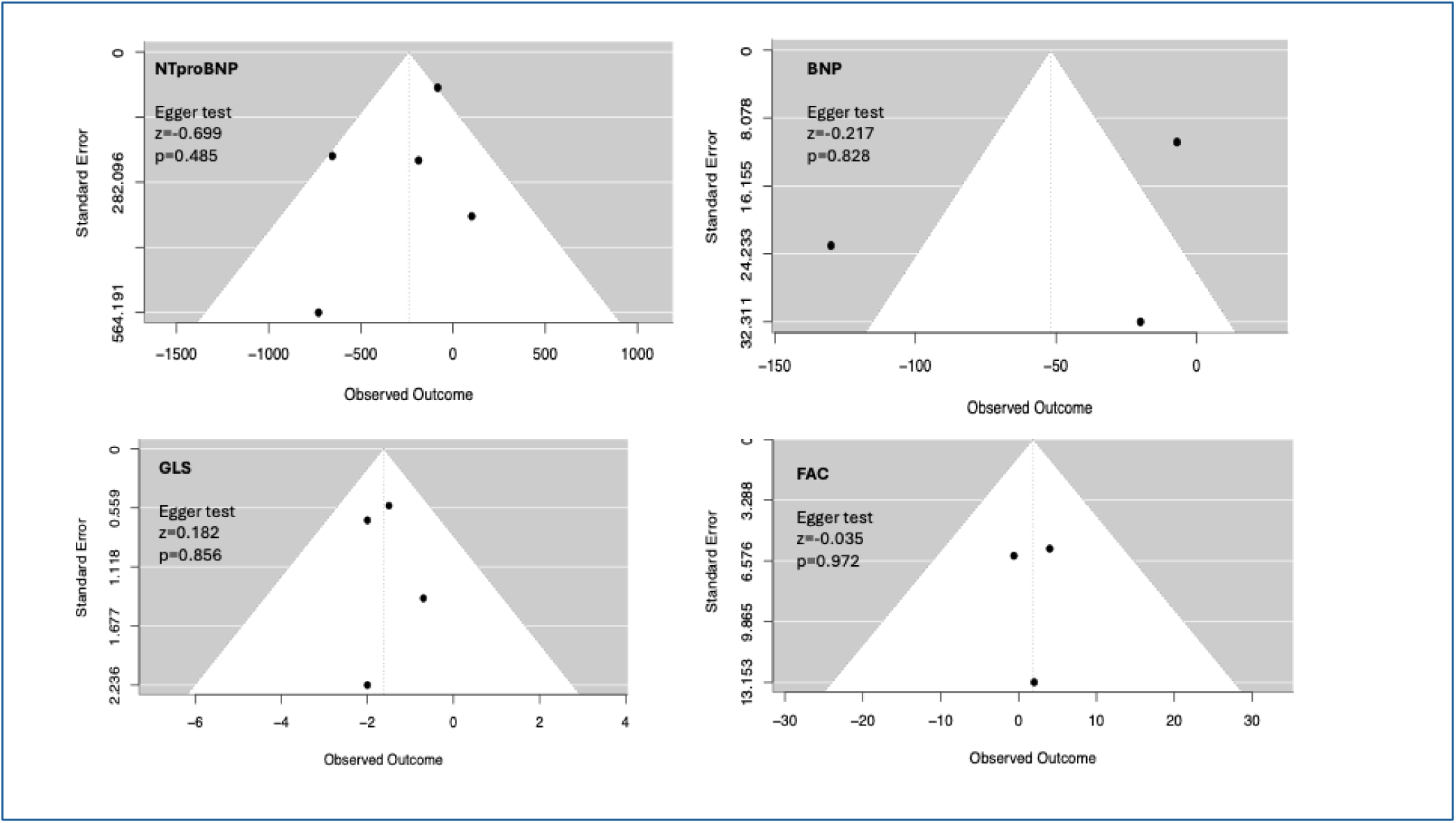
Funnel plots exploring risk of publications bias.

**Supplementary Table 1.** PRISMA Checklist.

|  | Item # | Checklist item | Location where item is reported |
| --- | --- | --- | --- |
| **TITLE** |  |  |  |
| Title | 1 | Identify the report as a systematic review. | 1 |
| **ABSTRACT** |  |  |  |
| Abstract | 2 | See the PRISMA 2020 for Abstracts checklist. | 2-3 |
| **INTRODUCTION** |  |  |  |
| Rationale | 3 | Describe the rationale for the review in the context of existing knowledge. | 10 |
| Objectives | 4 | Provide an explicit statement of the objective(s) or question(s) the review addresses. | 10 |
| **METHODS** |  |  |  |
| Eligibility criteria | 5 | Specify the inclusion and exclusion criteria for the review and how studies were grouped for the syntheses. | 11 |
| Information sources | 6 | Specify all databases, registers, websites, organisations, reference lists and other sources searched or consulted to identify studies. Specify the date when each source was last searched or consulted. | 11 |
| Search strategy | 7 | Present the full search strategies for all databases, registers and websites, including any filters and limits used. | Appendix |
| Selection process | 8 | Specify the methods used to decide whether a study met the inclusion criteria of the review, including how many reviewers screened each record and each report retrieved, whether they worked independently, and if applicable, details of automation tools used in the process. | 11, 14 |
| Data collection process | 9 | Specify the methods used to collect data from reports, including how many reviewers collected data from each report, whether they worked independently, any processes for obtaining or confirming data from study investigators, and if applicable, details of automation tools used in the process. | 11-12 |
| Data items | 10a | List and define all outcomes for which data were sought. Specify whether all results that were compatible with each outcome domain in each study were sought (e.g. for all measures, time points, analyses), and if not, the methods used to decide which results to collect. | 14-15 |
|  | 10b | List and define all other variables for which data were sought (e.g. participant and intervention characteristics, funding sources). Describe any assumptions made about any missing or unclear information. | 12 |
| Study risk of bias assessment | 11 | Specify the methods used to assess risk of bias in the included studies, including details of the tool(s) used, how many reviewers assessed each study and whether they worked independently, and if applicable, details of automation tools used in the process. | 13-14 |
| Effect measures | 12 | Specify for each outcome the effect measure(s) (e.g. risk ratio, mean difference) used in the synthesis or presentation of results. | 16 |
| Synthesis methods | 13a | Describe the processes used to decide which studies were eligible for each synthesis (e.g. tabulating the study intervention characteristics and comparing against the planned groups for each synthesis (item #5)). | n/a |
|  | 13b | Describe any methods required to prepare the data for presentation or synthesis, such as handling of missing summary statistics, or data conversions. | n/a |
|  | 13c | Describe any methods used to tabulate or visually display results of individual studies and syntheses. | 16 |
|  | 13d | Describe any methods used to synthesize results and provide a rationale for the choice(s). If meta-analysis was performed, describe the model(s), method(s) to identify the presence and extent of statistical heterogeneity, and software package(s) used. | 16 |
|  | 13e | Describe any methods used to explore possible causes of heterogeneity among study results (e.g. subgroup analysis, meta-regression). | n/a |
|  | 13f | Describe any sensitivity analyses conducted to assess robustness of the synthesized results. | n/a |
| Reporting bias assessment | 14 | Describe any methods used to assess risk of bias due to missing results in a synthesis (arising from reporting biases). | n/a |
| Certainty | 15 | Describe any methods used to assess certainty (or confidence) in the body of | n/a |
| assessment |  | evidence for an outcome. |  |
| **RESULTS** |  |  |  |
| Study selection | 16a | Describe the results of the search and selection process, from the number of records identified in the search to the number of studies included in the review, ideally using a flow diagram. | 16-17 |
|  | 16b | Cite studies that might appear to meet the inclusion criteria, but which were excluded, and explain why they were excluded. | 17 |
| Study characteristics | 17 | Cite each included study and present its characteristics. | 19-24 |
| Risk of bias in studies | 18 | Present assessments of risk of bias for each included study. | 30-31 |
| Results of individual studies | 19 | For all outcomes, present, for each study: (a) summary statistics for each group (where appropriate) and (b) an effect estimate and its precision (e.g. confidence/credible interval), ideally using structured tables or plots. | 27-28 |
| Results of syntheses | 20a | For each synthesis, briefly summarise the characteristics and risk of bias among contributing studies. | 25-31 |
|  | 20b | Present results of all statistical syntheses conducted. If meta-analysis was done, present for each the summary estimate and its precision (e.g. confidence/credible interval) and measures of statistical heterogeneity. If comparing groups, describe the direction of the effect. | 25-28 |
|  | 20c | Present results of all investigations of possible causes of heterogeneity among study results. | n/a |
|  | 20d | Present results of all sensitivity analyses conducted to assess the robustness of the synthesized results. | n/a |
| Reporting biases | 21 | Present assessments of risk of bias due to missing results (arising from reporting biases) for each synthesis assessed. | n/a |
| Certainty of evidence | 22 | Present assessments of certainty (or confidence) in the body of evidence for each outcome assessed. | 25-28 |
| **DISCUSSION** |  |  |  |
| Discussion | 23a | Provide a general interpretation of the results in the context of other evidence. | 32-39 |
|  | 23b | Discuss any limitations of the evidence included in the review. | 33-39 |
|  | 23c | Discuss any limitations of the review processes used. | 33-39 |
|  | 23d | Discuss implications of the results for practice, policy, and future research. | 39-40 |
| **OTHER INFORMATION** |  |  |  |
| Registration and protocol | 24a | Provide registration information for the review, including register name and registration number, or state that the review was not registered. | n/a |
|  | 24b | Indicate where the review protocol can be accessed, or state that a protocol was not prepared. | n/a |
|  | 24c | Describe and explain any amendments to information provided at registration or in the protocol. | n/a |
| Support | 25 | Describe sources of financial or non-financial support for the review, and the role of the funders or sponsors in the review. | 1 |
| Competing interests | 26 | Declare any competing interests of review authors. | 1 |
| Availability of data, code and other materials | 27 | Report which of the following are publicly available and where they can be found: template data collection forms; data extracted from included studies; data used for all analyses; analytic code; any other materials used in the review. | n/a |

